# OASIS-3: Longitudinal Neuroimaging, Clinical, and Cognitive Dataset for Normal Aging and Alzheimer Disease

**DOI:** 10.1101/2019.12.13.19014902

**Authors:** Pamela J. LaMontagne, Tammie LS. Benzinger, John C. Morris, Sarah Keefe, Russ Hornbeck, Chengjie Xiong, Elizabeth Grant, Jason Hassenstab, Krista Moulder, Andrei G. Vlassenko, Marcus E. Raichle, Carlos Cruchaga, Daniel Marcus

## Abstract

OASIS-3 is a compilation of MRI and PET imaging and related clinical data for 1098 participants who were collected across several ongoing studies in the Washington University Knight Alzheimer Disease Research Center over the course of 15 years. Participants include 605 cognitively normal adults and 493 individuals at various stages of cognitive decline ranging in age from 42 to 95 years. The OASIS-3 dataset contains over 2000 MR sessions, including multiple structural and functional sequences. PET metabolic and amyloid imaging includes over 1500 raw imaging scans and the accompanying post-processed files from the PET Unified Pipeline (PUP) are also available in OASIS-3. OASIS-3 also contains post-processed imaging data such as volumetric segmentations and PET analyses. Imaging data is accompanied by dementia and APOE status and longitudinal clinical and cognitive outcomes. OASIS-3 is available as an open access data set to the scientific community to answer questions related to healthy aging and dementia.

## BACKGROUND & SUMMARY

The Open Access Series of Imaging Studies (OASIS) is a multimodal collection of data focused on the effects of healthy aging and Alzheimer disease (AD) that is freely available to the scientific community. Previously released OASIS-Cross-sectional^1^ and OASIS-Longitudinal^2^ have been cited over 500 times. Data from these projects have been extensively used in the development of neuroimaging tools, such as brain atlases^3,4^, tissue segmentation algorithms^5^, and de-identification tools^6^, used to create models of healthy aging^7,8^ and AD timecourse^7–11^, used to answer hypothesis driven questions^12,13^, and used for educational and training purposes. Here we describe the OASIS-3 data release, which expands upon these prior releases.

OASIS-3 incorporates data from 1098 participants covering the adult life span aged 42 to 95, including cognitively normal individuals and individuals with early-stage AD dementia. The OASIS-3 release includes structural and functional MRI (magnetic resonance imaging), amyloid and metabolic PET (positron emission tomography) imaging, neuropsychological testing, and clinical data. Figure 1 shows the distribution of serial MRI and PET acquisition containing over 500 subjects with longitudinal MRI scans and over 400 with serial PET scans. The expansive data provided in OASIS-3 can be utilized as an individual dataset or in combination with other open data answering a multitude of research topics. Key features of OASIS-3 are:

**FIGURE 1.**
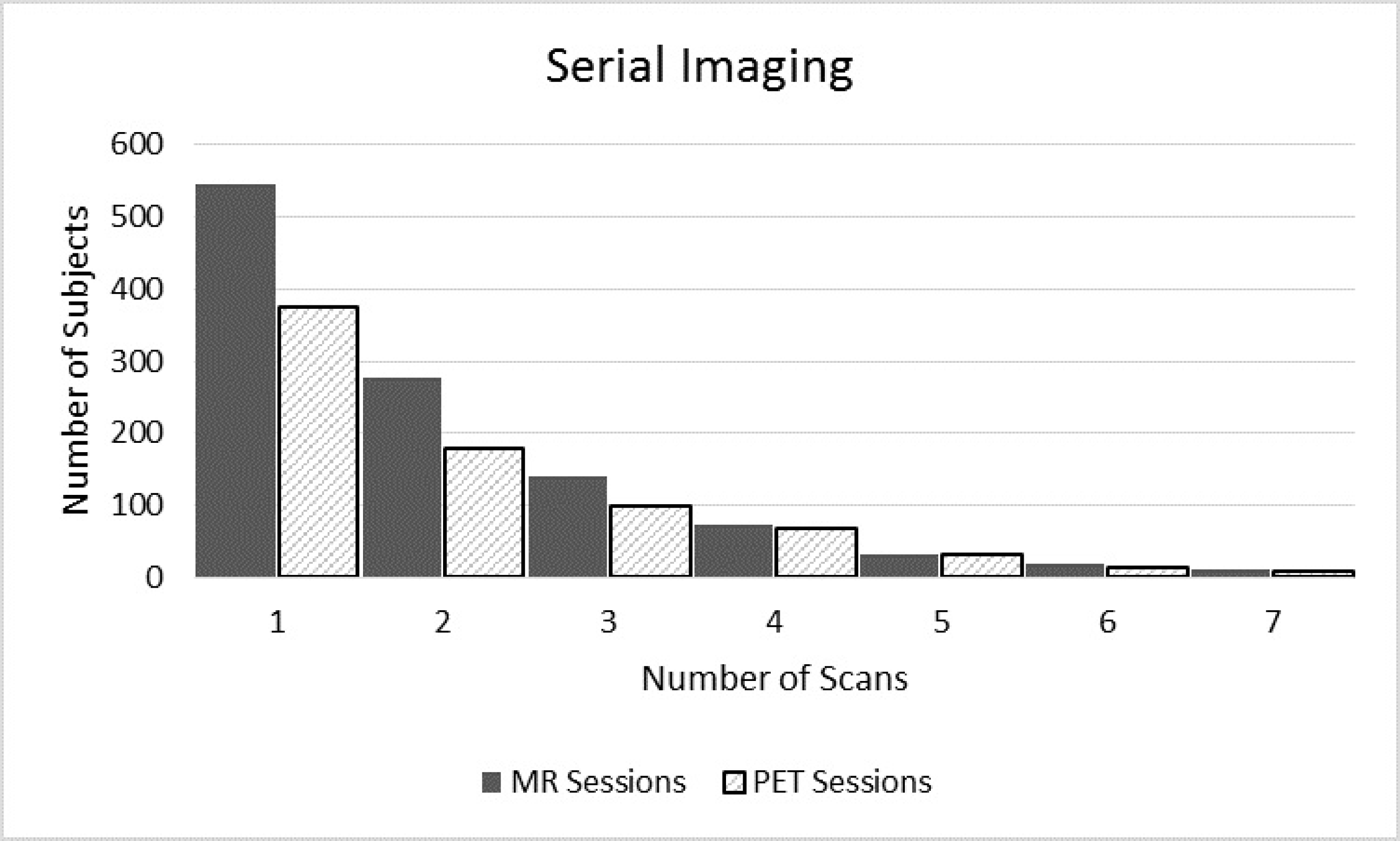
Serial Imaging. Distribution of MRI and PET sessions

1. Clinical and Cognitive assessments are standardized through the Uniform Dataset (UDS) allowing for combination with multiple Alzheimer open data projects
2. In addition to raw MRI and PET data, OASIS-3 includes post-processed statistical outputs, regional segmentations, and structure files. These can be used for a multitude of applications such as development of PET processing tools, improving volumetric segmentation, and analyzing individual structural differences.
3. OASIS-3 expands upon prior OASIS projects by providing longitudinal, multi- parametric MR imaging such as ASL, BOLD, DTI, SWI, and FLAIR. This variety of imaging data enables investigators to examine the longitudinal changes across multiple tissue types develop better imaging models of aging and cognitive impairment.

OASIS-3 is strong dataset that can be evaluated as an independent source or provide a valuable supplement to additional datasets. While there are other Alzheimer databases available, such as the Alzheimer’s disease Neuroimaging Initiative (ADNI), to the research community, OASIS-3 is unique in that the initial enrollment focused on pre- clinical cohort and followed longitudinal progression, whereas ADNI enrolled individuals with dementia or mild cognitive impairment. By providing Centiloid (Su et al^53^) data for all amyloid tracers OASIS-3 is helping with standardization in the neuroimaging community. Analyses of OASIS-3 data will aid in clinical trial design and provide pilot results and power calculations necessary for future funding. In the future we intend to provide updates to OASIS-3 with biomarker data (i.e. cerebrospinal fluid), tau PET imaging, and additional longitudinal timepoints. In summary, we are providing a dataset that will be a valuable asset to the scientific community.

## Methods

### Participants

#### Recruitment and Exclusion Criterion

OASIS-3 includes participants enrolled into several ongoing studies through the Charles F. and Joanne Knight Alzheimer Disease Research Center (Knight ADRC) at Washington University in St. Louis spanning over 15 years and several research studies - Memory and Aging Project, Adult Children Study^14^, and Healthy Aging and Senile Dementia. Each study targets varying cohorts but include similar assessments and visit intervals. The following cohorts were recruited: (1) Individuals who were generally healthy, were cognitively normal (CDR = 0), and had a family history of AD, defined as being a biologic child of at least one parent with a reported history of AD dementia with onset age 80 years; (2) individuals who were generally healthy, were cognitively normal (CDR = 0), and had no family history of AD for either biological parent and lived at least to age 70 years; and (3) healthy individuals 65 and older, both those who were cognitively normal (CDR 0) and this with very mild-mild symptomatic AD (CDR 0.5 and 1). Exclusion criterion included medical conditions that precluded longitudinal participation (eg, end-stage renal disease requiring dialysis) or medical contraindications for the study arms (e.g. pacemaker for MRI, anticoagulant use for lumbar puncture). Participants were recruited from the community via flyers, word of mouth, and community engagements. Participants from all cohorts agreed to submit an initial blood sample for genetic testing, complete regular cognitive testing, and neuroimaging and lumbar punctures approximately every 2-3 years. Each participant was enrolled along with a collateral source, someone who knew the participants well (eg, spouse of adult child) and could report whether the participant’s current cognitive and functional performance was or was not at previously attained levels. All participants were consented into Knight ADRC-related projects following procedures approved by the Institutional Review Board of Washington University School of Medicine.

#### Imaging inclusion/exclusion

Participants enrolled in studies at the Knight ADRC Clinical Core were referred to the Knight ADRC Research Imaging (KARI) Program for magnetic resonance imaging (MRI) and positron emission tomography (PET) scans. All participants were required to have a CDR ≤1 at the time of most recent Clinical Core assessment. Participants completed screening for general health information to assess any contraindications to PET or MR imaging. Participants were excluded for the following health reasons: women who were pregnant or breastfeeding; implanted medical devices such as pacemakers and drug pump; history or risk of metal in the eye; and history of claustrophobia. Eligible participants signed informed consent for one of the KARI imaging studies that included MR only, PET only, or MR and PET scans. To the best effort of investigators, participants underwent scan sessions within six months of Clinical Core visits. Across the years of scanning, gaps in funding, funding for additional sub-studies, or participant related delays, have caused variations in visit timelines resulting in extended or decreased intervals.

#### Clinical assessments and ADRC data collection procedures

Participants completed clinical assessment protocols in accordance with National Alzheimer Coordinating Center Uniform Data Set^15,16^ (UDS). UDS assessments included family history of AD, medical history, physical examination, and neurological evaluation. Participants age 64 or younger underwent clinical and cognitive assessments every 3 years. Participants age 65 and older had annual clinical and cognitive assessments. Dementia status was assessed for the UDS using the Clinical Dementia Rating^17^ (CDR) Scale, with CDR 0 indicating normal cognitive function, CDR 0.5 very mild impairment, CDR 1 mild impairment, and CDR 2 moderate dementia; once a participant reached CDR 2, they no longer were eligible for in-person assessments. The OASIS datatype “ADRC Clinical Data” includes age at entry, height, weight, CDR evaluations (UDS form B1 and B4 variables).

During the assessment, clinicians completed a diagnostic impression intake and interview culminating with a coded dementia diagnosis that is recorded in the OASIS datatype “ADRC Clinical Data” dx1-dx5. Diagnoses for this variable include “cognitively normal”, “AD dementia”, “vascular dementia” and contributing factors such as vitamin deficiency, alcoholism, and mood disorders. The diagnostic determination for variables dx1-dx5 is separate from UDS assessments, however they may overlap in diagnostic conclusions.

UDS form A1 includes participant demographics such as disease status at study entry, sex, race, language, education, type of residence, level of independence, marital status, and handedness. UDS form A2 includes demographic information about the participant’ designated informant. Information about the AD status of the participant’s parents, siblings, children, and other relatives is available in UDS form A3. Participant’s health history is reported in UDS form A5. Cerebrovascular health and stroke history are recorded in UDS form B2. UDS form B3 represents the Unified Parkinson’s Disease Rating Scale (UPDRS). Form B5 records the participant’s neuropsychiatric history while Form B6 includes the Geriatric Depression Scale. UDS Form B7 includes the Functional Assessment Questionnaire including questions about daily activities. Clinician reports include Form B8 - Physical and neurological findings, Form B9 - clinician based judgement of symptoms, and Form D1 – Clinician diagnosis – Cognitive status and Dementia.

#### Neuropsychological Assessment

Participants aged 65 and older annually completed a battery of neuropsychological tests at the ADRC^16,18^. The Mini-Mental State Examination (MMSE) measures general cognitive status, with scores ranging from 0 (severe impairment to 30 (no impairment). Logical Memory - Story A, a subtest of the Wechsler Memory Scale-Revised measures episodic memory. Participants recall as many details as they can from a short story containing 25 bits of information after it is read aloud by the examiner and again after a 30-minute delay, with scores ranging from 0 (no recall) to 25 (complete recall). Digit Span requires participants to repeat back a series of digits forward and backwards providing measures of attention and working memory. Scores were based on the number of trials repeated correctly forwards and backwards, as well as the longest length the participant is able to repeat back. Semantic memory and language were measured using the Category Fluency test, requiring participants to name as many words belonging to a category, animal and vegetable, in 60 seconds, and the Boston Naming Test, in which participants name drawings of common objects. Psychomotor speed was measured using the WAIS-R Digit Symbol test and the Trail Making Test Part A. The WAIS-R Digit Symbol test is scored on the number of digit symbol pairs completed in 90 seconds. Executive function was measured using the Trail Making Test Part B. In the Trail Making Test participants were asked to make a trail by connecting a series of numbers (1-26) for part A and connecting a series of alternating numbers and letters (1-A-2-B) for part B. Outcome measures include total time to complete in seconds with a max of 150s for Trails A and 300s for Trails B, number of commission errors, and number of correct lines. These results can be found in the UDS Form C1 reports in the Psych Assessment form.

#### Genotyping (APOE)

The apolipoprotein E gene (APOE) has been identified for its influence on Alzheimer’s disease. The APOE ε4 allele of apolipoprotein E gene (APOE ε4) has been linked to increased risk for Alzheimer’s disease^19^ while the ε2 allele (APOE ε2) may provide protection from Alzheimer’s disease^20,21^. Genomic DNA was extracted from blood samples using QIAmp DNA blood mini kits from Qiagen Inc. (Valencia, CA). APOE genotyping was performed using PCR amplification of a 244-bp fragment followed by restriction enzyme HhaI digest^22^.

### Neuroimaging Data description

#### Scanners

All neuroimaging scans were conducted by the Knight Alzheimer Research Imaging Program at Washington University in St. Louis. MRI was collected on 3 different Siemens scanner models (Siemens Medical Solutions USA, Inc): Vision 1.5T, TIM Trio 3T (2 different scanners of this model), and BioGraph mMR PET-MR 3T. Positron emission tomography (PET) was conducted on 3 different Siemens PET scanners: ECAT HR+ 962 PET scanner, Biograph 40 PET/CT scanner, and BioGraph mMR PET-MR 3T. All sessions collected simultaneously on the BioGraph mMR scanner have been split into individual PET and MRI sessions for archiving purposes. Participants were placed in a 16-channel head coil on 1.5T scanners and 20-channel head coil on 3T scanners with foam pad stabilizers placed next to the ears to decrease motion. Some participants had a vitamin-E fiducial marker placed on the left temple.

### MRI

#### MRI Sequences

MR imaging includes various anatomical and functional sequences. High resolution structural sequences include T1-weighted, T2-weighted, FLAIR, and TSE, susceptibility weighted imaging (SWI), diffusion weighted imaging (DWI), arterial spin labeling (ASL), resting state BOLD, fieldmaps, time of flight (TOF). Susceptibility weighted imaging (SWI) sequences include magnitude, phase, minimum intensity projection (mIP), and a combination of the magnitude and phase creating a single SWI image. SWI imaging is used to identify micro hemorrhages. Arterial spin labeling (ASL) is a non-invasive sequence designed to evaluate cerebral blood flow similar to invasive perfusion sequences. Time of Flight (TOF) is an angiography sequence designed to look at blood vessels without administration of a contrast agent. Participants were asked to lay quietly with their eyes open while two 6 minute resting state BOLD sequences were collected to evaluate neural activity at rest. Diffusion tensor imaging (DTI) sequences included in OASIS-3 were designed to investigate fractional anisotropy, apparent diffusion coefficient, and can be used to create 3D white matter tracts. Diffusion sequences collected on the PET-MR were acquired with 64 directions, while sequences collected on other scanner models were acquired with 24-26 directions. Fieldmap images were acquired to correct for field inhomogeneities and often used with BOLD and DTI imaging. Acquisition parameters for each sequence are available in the accompanying JSON (JavaScript Object Notation) file. A spreadsheet is provided with the OASIS-3 data including common parameters of interest for each scan type.

#### MRI Post-Processing: FreeSurfer volumetric segmentation

All MR imaging sessions were subjected to cortical reconstruction and volumetric segmentation of T1-weighted images using the FreeSurfer image analysis suite, which is documented and freely available for download online (http://surfer.nmr.mgh.harvard.edu/). The technical details of these procedures are described in prior publications^23–34^. Briefly, this processing includes motion correction and averaging^34^ of volumetric T1 weighted images, removal of non-brain tissue using a hybrid watershed/surface deformation procedure^33^, automated Talairach transformation, segmentation of the subcortical white matter and deep gray matter volumetric structures (including hippocampus, amygdala, caudate, putamen, ventricles)^26,27^ intensity normalization^35^, tessellation of the gray matter white matter boundary, automated topology correction^25,36^, and surface deformation following intensity gradients to optimally place the gray/white and gray/cerebrospinal fluid borders at the location where the greatest shift in intensity defines the transition to the other tissue class^23,24^.

Once the cortical models are complete, a number of deformable procedures were performed for further data processing and analysis including surface inflation (Fischl et al., 1999a), registration to a spherical atlas which is based on individual cortical folding patterns to match cortical geometry across subjects (Fischl et al., 1999b), parcellation of the cerebral cortex into units with respect to gyral and sulcal structure^37^ (Fischl et al., 2004b), and creation of a variety of surface based data including maps of curvature and sulcal depth. This method uses both intensity and continuity information from the entire three dimensional MR volume in segmentation and deformation procedures to produce representations of cortical thickness, calculated as the closest distance from the gray/white boundary to the gray/CSF boundary at each vertex on the tessellated surface^24^. The maps are created using spatial intensity gradients across tissue classes and are therefore not simply reliant on absolute signal intensity. The maps produced are not restricted to the voxel resolution of the original data thus are capable of detecting submillimeter differences between groups. Procedures for the measurement of cortical thickness have been validated against histological analysis^38^ and manual measurements^39,40^. FreeSurfer morphometric procedures have been demonstrated to show good test-retest reliability across scanner manufacturers and across field strengths^31,41^.

Data included in OASIS-3 were processed using an XNAT pipeline for the FreeSurfer image analysis suite using Dell PowerEdge 1950 servers with Intel Xeon processors running CentOS 5.5 Linux. T1-weighted images collected on 1.5T scanners were processed with FreeSurfer v5.0 or v5.1 with the patch correcting for inaccurate ICV values (FreeSurfer Patch 10Dec2012). T1-weighted images from the 3T scanners were segmented using FreeSurfer 5.3 including the 2012 patch (FreeSurfer Patch 10Dec2012) and the Human Connectome (HCP) patch (ftp://surfer.nmr.mgh.harvard.edu/pub/dist/freesurfer/5.3.0-HCP) designed to correct pial surface mapping. Following segmentation, a trained lab member reviewed the images for accurate segmentation and assigned a quality control status. If segmentation did not pass quality control, then it underwent edits using TkMedit (http://freesurfer.net/fswiki/TkMedit), a FreeSurfer toolbox, and the revised images were rerun through the XNAT pipeline. Upon completion the segmentations were reviewed again and assigned a status of “passed with edits” or “fail”. OASIS-3 includes the regional volume statistics and the post-processed output files for all segmentations that passed QC. Segmentations with a “fail” QC status were excluded from OASIS-3. Statistical data for volumetric regions is available for download in spreadsheet format. FreeSurfer post-processed output images such as surface maps and segmented volumes have also been provided for download.

### PET

#### Tracers and Acquisition Protocols

Participants underwent positron emission tomography on one of three different Siemens scanners: ECAT HRplus 962 PET scanner, Biograph 40 PET/CT scanner, Biograph mMR PET-MR. A softened thermoplastic mask with enlarged eye holes was placed over the head and secured to minimize head motion or HRplus and PET/CT scanners. A transmission scan was obtained for attenuation correction on the HRplus scanner. On the Siemens Biograph 40 PET/CT scanner a three-second X-ray Topogram was acquired in the lateral plane to visualize the head and determine the exact scan position. Prior to the PET scan, a spiral CT (Computed Tomography) scan was performed for attenuation correction at the low dose CT. On the Siemens mMR PET-MR a series of Dixon based tissue segmented MR scans were acquired to create a Dixon μmap used for attenuation correction. Frame timing information for each scan is provided in an accompanying tsv file.

#### β-amyloid (Aβ) Imaging

Two radiopharmaceuticals, Pittsburgh Compound B ([^11^C]PIB or PIB) and Florbetapir [^18^F] (^18^F-AV-45 or AV45), were used to investigate β-amyloid (Aβ) deposits in the brain. For PIB PET scans, participants received an I.V. bolus administration of 6 - 20 mCi of [^11^C]PIB with a 60 minute dynamic PET scan in 3D mode (24 ⨯ 5 sec frames; 9 ⨯ 20 sec frames; 10 ⨯ 1 min frames; 9 ⨯ 5 min frames). For Florbetapir PET scans, participants received a single I.V. bolus administration of 10 mCi (+-10%) of [^18^F]AV45 with dynamic PET scan in 3D mode. There were two acceptable procedures for obtaining Florbetapir PET scans to capture the 50-70min uptake window: (1) in the preferred approach, the participants were positioned in the PET-MR scanner at the time of injection and a dynamic 70-minute scan (consisting of 4 ⨯ 15sec frames, 8 ⨯ 30sec frames, 9 ⨯ 1 min, 2 ⨯ 3 min frames, 10 ⨯ 5 min frames) was obtained starting at the time of injection (2) for those participants who were unable to tolerate the full exam, PET-MR scanning began 50 minutes after Florbetapir injection with a dynamic 20min (consisting of 4×5min frames) PET acquisition. In the event of technical problems participants may have been scanned on an alternate available PET scanner. All PET scan parameters are available in the accompanying JSON file.

#### Metabolic Imaging

Metabolic imaging with FDG was performed on the HR+ PET scanner. Participants were asked to eat a normal breakfast then fast for 4hrs prior to FDG administration. Prior to tracer administration plasma glucose concentration was verified and individuals were excluded if blood glucose concentrations >=165. Participants received an I.V. bolus injection of 5 mCi of [^18^F]FDG followed by a dynamic 60min (consisting of 24 ⨯ 5 sec frames, 9 ⨯ 20 sec frames, 10 ⨯ 1 min frames and 9 × 5 min frames) PET acquisition.

#### PET Post-Processing: PUP

PET imaging analyses were performed using the PET unified pipeline^42,43^ (PUP, https://github.com/ysu001/PUP) via XNAT on Dell PowerEdge 1950 servers with Intel Xeon processors running CentOS 5.5 Linux. PET images were smoothed to achieve a common spatial resolution of 8mm to minimize inter-scanner differences^44^. Inter-frame motion correction for the dynamic PET images is performed using standard image registration techniques^45,46^. PET to MR registration is performed using a vector-gradient algorithm^47^ (VGM) in a symmetric fashion (i.e. average transformation for PET->MR and inverse of MR->PET was used as the final transformation matrix). Regional PET processing is performed based on FreeSurfer segmentation using wmparc.mgz as the region definition, and each FreeSurfer region is analyzed. When FreeSurfer processing of matching T1w image failed, PET processing was excluded from OASIS-3. The PUP pipeline generates both reports of regional measurements as well as a voxel-wise SUVR image in the individual FreeSurfer space.

PUP also accounts for partial volume effects (PVE) that are the result of spatial distortion from low resolution PET images. The distortion caused by PVE is directly related to the size and shape of the region of interest in addition to spatial resolution of the images. In longitudinal studies, the impact of PVE can be further confounded by brain atrophy due to aging and pathological changes. PUP accounts for these distortion correction technique is implemented using a regional spread function (RSF) based approach^42,48^. Su et al.^42,49^ has demonstrated that the RSF technique was able to improve PET quantification and achieve better sensitivity to longitudinal changes in amyloid burden. PUP processed results include results both with and without RSF partial volume correction and SUVR voxel-wise images produced without partial volume correction.

Two modeling approaches are implemented in PUP using the cerebellum as a reference region. Binding potential (BPND) is calculated using Logan graphical analysis^42,43,49–51^, for full dynamic PET imaging data are available, i.e. PET acquisition was started in synchronization with tracer administration and PET images were reconstructed into multiple time frames. Non-logan graphical analysis is used to process scans that do not include full dynamic PET imaging but no binding potential is calculated. Regional target-to-reference intensity ratio, a.k.a, standard uptake ratio (SUVR), is estimated for all PET data. PET analysis (both BPND and SUVR) used peak time windows for each tracer, 30 to 60 minutes post-injection for PiB, 50 to 70 minutes for Florbetapir, and 40 to 60 minutes for FDG.

#### PET Post-Processing: Centiloid Standardization

Amyloid specific processing assessed global amyloid burden of each tracer and included a Centiloid conversion to provide comparable values across amyloid tracers. To assess global amyloid burden based on amyloid PET imaging data, the mean cortical binding potential (MCBP) or mean cortical SUVR (MCSUVR) were calculated from the arithmetic mean of BPND or SUVRs from four critical regions: 1) precuneus (PREC) defined as the combined left and right hemisphere ctx-precuneus, 2) prefrontal cortex (PREF) defined as the left and right combined ctx-superior frontal and ctx-rostral middle frontal regions, 3) gyrus rectus (GR) defined as the left and right combined ctx- lateral orbitofrontal and ctx-medial orbitofrontal regions, and 4) lateral temporal (TEMP) defined as the left and right combined ctx-superior temporal and ctx-middle temporal regions^43^. In order to provide standard quantification of amyloid analysis across lab protocols and tracers, the Centiloid scale has been developed to convert amyloid PET data to a 0-100 scale^52^. Both PIB and Florbetapir amyloid imaging have been calibrated on the Centiloid scale based on equations presented in Su et al^53^ and are available in OASIS-3.

### Data Records

#### OASIS-3 data inclusion/exclusion

In order to provide standardized clinical and cognitive measures, data is only included in the OASIS-3 database if participants had at least one imaging session after the implementation of the Uniform Data Set^15,16^ (UDS) in 2005. Imaging scans collected prior to the implementation of the UDS may be included for eligible participants. Any imaging sessions without accompanying UDS assessments were excluded. Post-processed data that did not pass quality control review (described in the image processing section) were excluded from OASIS-3. Participants consented to the use of their data by the scientific community and data sharing terms have been approved by the Washington University Human Research Protection Office.

#### Participant description

OASIS-3 includes 1098 subjects ranging in age 42.5-95.6 yrs. The cohort is 84% Caucasian and 15% African American, with five individuals reporting Hispanic ethnicity. Table 1 provides a detailed description of study demographics. Demographic information available in OASIS-3 includes sex, age, handedness, race, ethnicity, socio- economic status, and marital status. 850 individuals entered with a baseline Clinical Dementia Rating Scale^17^ (CDR) of 0; 605 of those remained cognitively normal while 245 converted to cognitive impairment. An additional 248 participants were CDR > 0 at their baseline visit (Table 2). The APOE ε4 allele was present in 439 participants.

**Table 1.** Subject Demographics.

|  | **Male** | **Female** | **Total** |
| --- | --- | --- | --- |
| **N** | 487 | 611 | 1098 |
| **Age** | 70.17 (42.5-91.7) | 67.78 (43.2-95.6) | 68.84 (42.5-95.6) |
| **Right Handed** | 433 | 546 | 979 |
| **APOE** |  |  |  |
| **22** | 5 | 3 | 8 |
| **23** | 52 | 63 | 115 |
| **24** | 16 | 22 | 38 |
| **33** | 225 | 284 | 509 |
| **34** | 157 | 185 | 342 |
| **44** | 21 | 38 | 59 |
| **Race** |  |  |  |
| **Caucasian** | 428 | 498 | 926 |
| **African-American** | 57 | 110 | 168 |
| **Asian** | 2 | 3 | 5 |

**Table 2.** Clinical Dementia Rating (CDR) Distribution.

|  | *max CDR* |  |  |  |  |
| --- | --- | --- | --- | --- | --- |
| *min CDR* | **0** | **0.5** | **1** | **2 >** | **Total** |
| **0** | 605* | 192 | 39 | 14 | 850 |
| **0.5** |  | 66 | 61 | 52 | 179 |
| **1>** |  |  | 31 | 38 | 69 |
| **Grand Total** | 605 | 258 | 131 | 100 | 1098 |

\*Healthy controls

#### Data description

A total of 2168 MR sessions (236 1.5T and 1932 3T) are included in OASIS-3 with a variety of scan types to examine structure, vasculature integrity, and functional networks. Table 3 provides a detailed inventory of the scan types by scanner strength. T1w images were used for volumetric segmentation via FreeSurfer, and 1912 segmentations passed quality control review and have been included in OASIS-3. The FreeSurfer segmentations were used to calculate whole brain, total cerebral cortex, cortical white matter, and subcortical gray volumes provided with the OASIS-3 release. OASIS-3 includes 1608 PET imaging sessions comprised of 999 PiB, 492 Florbetapir (AV45), and 117 FDG scans (Table 4). Quality control reviewed post-processing PUP output^42,43^, which includes uptake measure for FreeSurfer-based regions, was included for 1356 PET sessions. Centiloid values^53^ have been provided to standardize values across amyloid tracers PiB and Florbetapir.

**Table 3.** MRI Scan type counts.

| **Scan Type** | **1.5T MR Sessions** | **3.0T MR Sessions** | **Total # of MR Session** |
| --- | --- | --- | --- |
| T1w | 236 | 1929 | 2165 |
| T2w | 230 | 1755 | 1985 |
| FLAIR | 0 | 735 | 735 |
| DTI | 0 | 1472 | 1472 |
| Bold – Resting State | 2 | 1689 | 1691 |
| ASL | 0 | 722 | 722 |
| SWI | 2 | 1217 | 1219 |
| TOF | 1 | 557 | 558 |
| Fieldmap | 2 | 977 | 979 |

**TABLE 4.** PET Scan type counts.

| **Tracer** | **Count** | **PUP** |
| --- | --- | --- |
| PIB | 1000 | 937 |
| AV45 | 492 | 419 |
| FDG | 117 | 0 |

#### Data Standardization and Anonymization Procedures

All participants were given unique OASIS-3 identifiers (ex. OAS3####). In compliance with HIPAA regulations, Protected Health Information (PHI) has been excluded from the release. Individuals aged 90> were able to be included by removing all referential dates from the released data we were able to include data for individuals aged 90>. Additional altered variables include: original participant identifiers, free text fields, location specific information stored in supplemental files. Across all datatypes, dates and visits are now represented as days from entry into the parent Knight ADRC project.

Because OASIS-3 images were acquired over 10+ years on multiple scanners, source images were generated in a diversity of file formats (DICOM, IMA, ECAT). To provide a single standard format, the sources data files were converted to NifTI format files using the dcm2niix^54^ conversion program and are organized and named following the Brain Imaging Data Structure^55^ (BIDS). One benefit of using BIDS is that the supplied NifTI files include only geometric information about the image and, therefore, free of protected health information. A second benefit of adopting the BIDS data structure is that it provides a standardized naming convention, file organization, and supplemental file information that can be used to facilitate downstream processing and analysis. Key acquisition information, such as magnet strength, scanner model, repetition time, and slice timing typically found in the DICOM formatted data are provided with the BIDS supplemental metadata in a JSON file. Raw and processed data, such as FreeSurfer and PUP outputs and statistics, underwent rigorous de-identification procedures that removed original subject IDs, scan IDs, and procedure dates from images and statistics files and replaced them with OASIS labels and generic dates (ex. 01/01/1970) or days from entry (ex. d1234). These methods were intended to remove all protected health information while maintaining provenance.

#### Data Sharing and Access

OASIS-3 data is openly available to the scientific community at https://www.oasis-brains.org. Data sharing was granted by participants through informed consent. Prior to accessing the data, users are required to agree to the OASIS Data Use Terms (DUT), which follow the Creative Commons Attribution 4.0 license. These terms require the user to acknowledge the use of OASIS data and data derived from OASIS data when publicly presenting any results or algorithms that benefitted from their use. Users are also expressly forbidden from attempting to identify participants. Once a user agrees to the DUT, they are provided with login credentials. The OASIS-3 data are hosted on XNAT Central^56^ (https://central.xnat.org), a publicly accessible data-repository. Each data point on XNAT Central has a unique identifier with URL that provides a persistent link to the data.

## Technical Validation

### Exploratory MRI and PET Analyses

Exploratory descriptive analyses were performed on the MRI and PET data in OASIS-3 to demonstrate feasibility of MRI and PET analyses in conjunction with behavioral measures. Groups were defined based on their longitudinal CDR score - Stable Controls as individuals with CDR = 0; Converters started with CDR = 0 and progressed to CDR >0; Dementia at aging started in the study with CDR > 0. Figure 2a depicts the longitudinal trajectory of whole brain volume (WBV) atrophy in stable controls compared to converters and those with dementia at entry. There is a significant difference in baseline WBV (*F(2,974)* = 32.36, *p* < 0.001; Figure 3a) with larger baseline WBV in both stable controls (*t*(749) = 7.98, *p* < 0.001) and converters (*t*(424) = 5.77, p < 0.001) compared to those with dementia but no difference between those scanned at CDR=0 (*t*(771) = 1.54, *p* = 0.125). Annual atrophy rates were calculated by subtracting the baseline WBV from final scan WBV, dividing by number of years for this interval, and multiplying by 100. Longitudinal analysis showed a group effect (*F*(2,501) = 8.06, *p* < 0.001, Figure 3b) with greater atrophy rates in those that entered with dementia (-2.01% per year, SD = 2.15) and converters (-1.68% per year, SD = 1.91) compared to stable controls (-1.13% per year, SD = 1.49; respectively (*t*(469) = 3.32, *p* < 0.001; (*t*(370) = 3.02, *p* = 0.002)) but no difference between those who had dementia at follow-up visits (*t*(159) = 0.829, *p* = 0.41).

**Figure 2.**
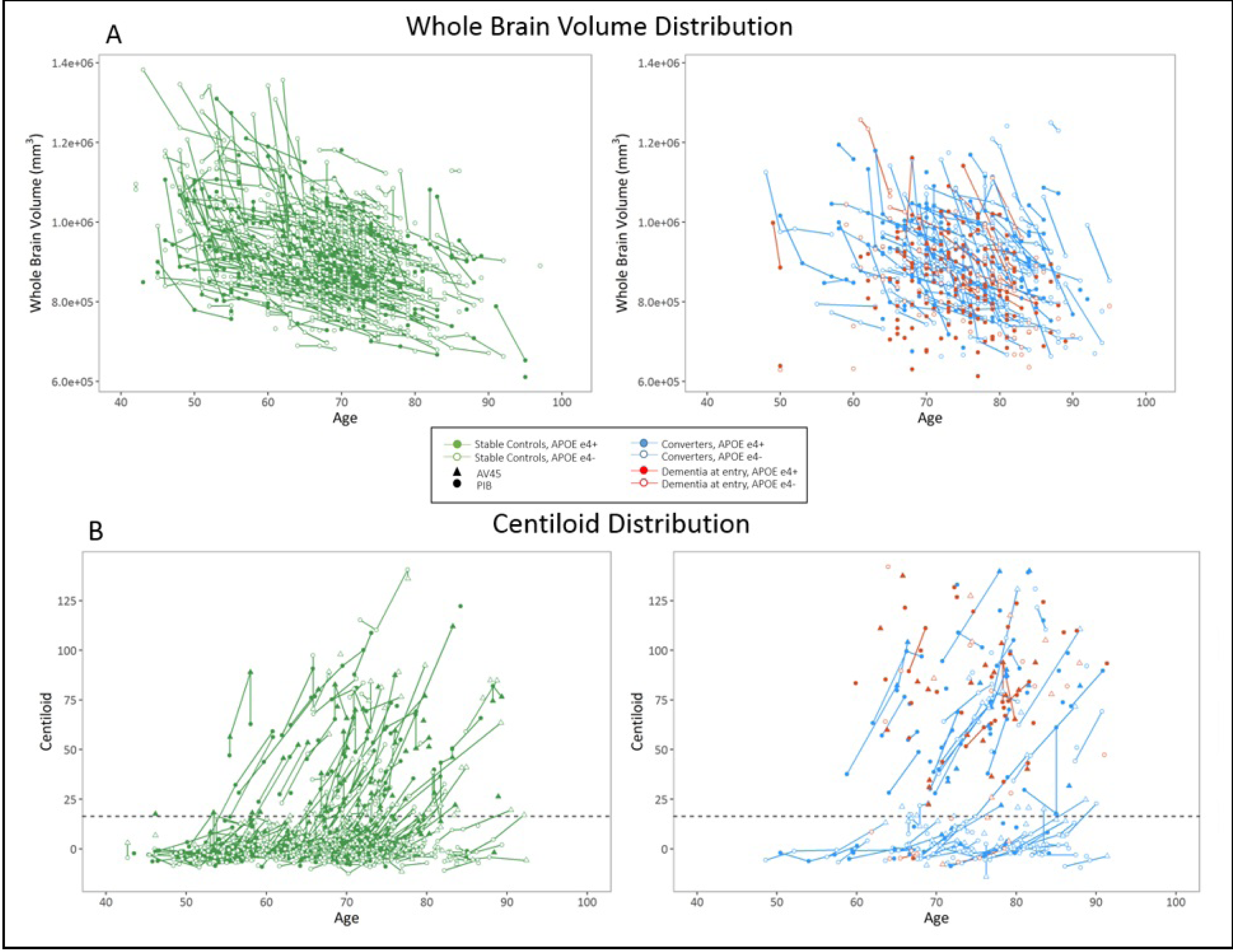
Imaging features in cognitively normal and demented adults in OASIS-3. A) Whole Brain Volume in healthy controls and individulas who had dementia at study entry or converted to dementia. APOE 4 allele carriers are indicated with --- line. B) Centiloid values (mSUVR) for healthy controls and individulas who had dementia at study entry or converted to dementia. Tracer at each timepoint is indicated with a triangle for AV45 and a circle for PIB. APOE 4 allele carriers are indicated with --- line.

**Figure 3.**
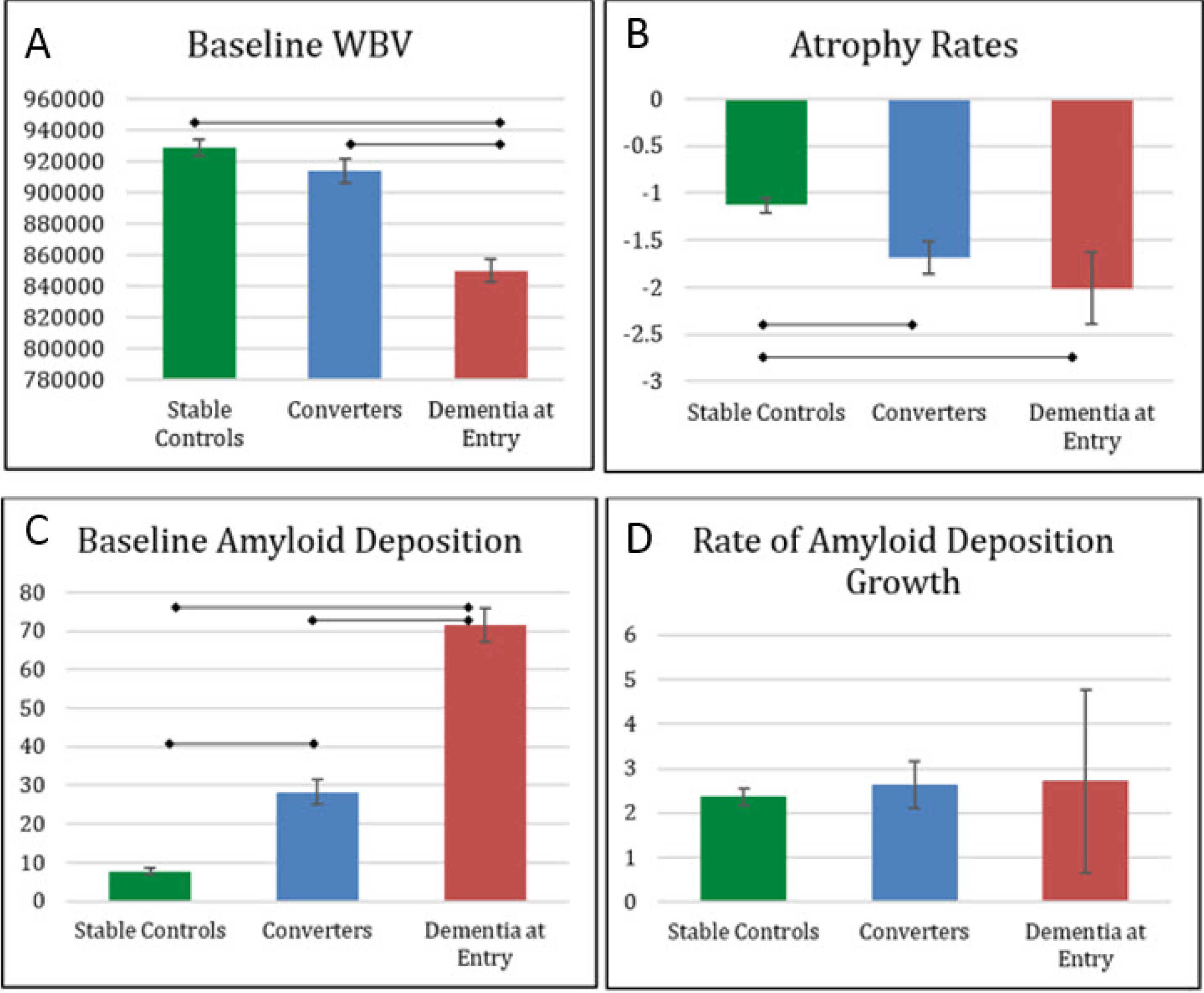
Baseline and Longitudinal patterns of Whole Brain Volume and Amyloid Deposition. Whole Brain Volume (a) at baseline scans and (b) percent annual WBV atrophy rates. Partial whole brain corrected Centiloid values for Amyloid deposition (c) at baseline scans and (d) percent annual deoposition rates.

In PET imaging, PIB and Florbetapir (AV45) amyloid tracers have shown greater cortical uptake in patients diagnosed with AD compared with control participants^11,57^. We conducted a cursory review of amyloid deposition levels in each group using partial volume corrected Centiloid values from PET scans. The longitudinal distribution of amyloid deposition is presented in Figure 2b with amyloid positivity marked at a Centiloid value of 16.4. As expected, the groups are significantly different (*F*(2,724) = 191.39 ; *p* < 0.001, Figure 3c) at the baseline scan. Those who entered the study with dementia (defined as CDR > 0.5) had significantly higher amyloid levels than those who later converted (*t*(229) = -8.08, *p* < 0.001) and stable controls (*t*(573) = -21.98, *p* < 0.001). Converters also had more amyloid as baseline scan compared to stable controls (*t*(646) = -8.46, *p* < 0.001). Rate of amyloid deposition was calculated by dividing the difference in baseline and last amyloid values by years to last follow-up PET scan. There was no significant difference in rate of amyloid deposition for all three groups (*F*(2,330) = 0.19 ; *p* > 0.05, Figure 3d).

OASIS-3 analyses show baseline amyloid deposition is correlated with later conversion to dementia but baseline whole brain volume is not significantly related to later conversion. The rate of change, however, is important for WBV but not for amyloid deposition. The pattern of WBV change indicates that converters have WBV similar to healthy controls however they have a steeper rate of decline and those that have dementia at baseline have a steeper decline of WBV loss. This indicates that WBV loss is a significant predictor of dementia in addition to the traditional analyses of smaller volumes such as the hippocampal and parietal cortices. An important finding shows that increased amyloid deposition levels precede measurable cognitive and clinical declines in preclinical AD population. The early deposition patterns and atrophy rates support early biomarker detection with intervention prior to observable declines in addition to longitudinal follow-up for volume changes.

These exploratory analyses are representative of simple analyses that can be conducted with the OASIS-3 data to describe factors related to cognitive decline. However, multimodal models provide a more comprehensive evaluation of the interaction of variables such as combining WBV atrophy rates and amyloid deposition rates to predict cognitive decline patterns^11,58–61^. We provide OASIS-3 data for investigators to explore changes across domains using alternate image processing methods and advanced statistical approaches in an effort to predict healthy aging and Alzheimer’s disease progression.

### Data Usage

A standardized visit structure was not maintained across the duration of the data collection. Investigators are instructed to use “days from entry” or “age at visit” to link imaging and clinical visits. In order to provide the most complete dataset we did not impose an association between visits. We have found that investigators vary on criterion used to combine imaging and assessments into a visit, i.e. clinical assessment within 6 months of imaging visit. Additionally, there is no categorical variable that indicates if an individual is healthy or has Alzheimer Disease as the investigators may choose to use various biomarkers markers to identify early stages of decline before an individual is symptomatic.

The published OASIS datasets – OASIS Cross-sectional, OASIS Longitudinal, and OASIS-3 – contain overlapping participant data and should never be combined or aggregated. It is recommended that investigators carefully consider which OASIS data set best fits their needs and limit their research to that data set. Due to mandatory anonymization procedures, links to subject data across datasets cannot be provided and users of OASIS data are prohibited from attempting to link OASIS data.

## Data Availability

OASIS-3 data is openly available to the scientific community at https://www.oasis-brains.org. Prior to accessing the data, users are required to agree to the OASIS Data Use Terms.

## Acknowledgements

Funding for the Knight ADRC and KARI were provided by NIH P50AG00561, P30NS09857781, P01AG026276, P01AG003991, R01AG043434, R01AG054567, UL1TR000448, and R01EB009352. Florbetapir doses were provided by Avid Radiopharmaceuticals, a wholly owned subsidiary of Eli Lilly.

## Author Contributions

Experimental Design: T.L.S.B., J.C.M., E.G., C.X., J.H., K.M., A.G.V., M.E.R., C.C., D.M. Data Curation: P.J.L., S.K., E.G., R.H. Drafting of the Manuscript: P.J.L., T.L.S.B., D.M. All authors provided critical review and editing of the manuscript.

## Abbreviations

AD: Alzheimer Disease
ADRC: Alzheimer Disease Research Center
APOE: 
ASL: arterial spin labeling
AV45: Florbetapir (^18^F-AV-45)
BPND: binding potential
CDR: Clinical Dementia Rating
CSF: cerebrospinal fluid
DWI: diffusion weighted imaging
DAT: is outmoded; use “AD dementia” or “symptomatic AD” – the latter term can be stated to encompass both MCI due to AD and AD dementia
FDG: fluorodeoxyglucose (^18^F-FDG)
BOLD: blood oxygen level dependent
MCBP: mean cortical binding potential
mCi: millicurie
MCSUVR: mean cortical SUVR
MRI: magnetic resonance imaging
OASIS: Open Access Series of Imaging Studies
PiB: Pittsburgh compound B (^11^C-PIB)
PUP: Pet Unified Pipeline
PET: Positron emission tomography
PVE: partial volume effects
QC: quality control
rsf: regional spread function
SWI: susceptibility weighted imaging
SUVR: standard uptake ratio
TOF: time of flight
UDS: Uniform Data Set

